# Continuous positive airway pressure but not Liraglutide-mediated weight loss improves early cardiovascular disease in obstructive sleep apnea: Data from a randomized proof-of-concept study

**DOI:** 10.1101/2023.05.23.23290424

**Authors:** Cliona O’Donnell, Shane Crilly, Anne O’Mahony, Brian O’Riordan, Mark Traynor, Rachael Gitau, Kenneth McDonald, Mark Ledwidge, Donal O’Shea, David J. Murphy, Jonathan D. Dodd, Silke Ryan

## Abstract

**Background:** Obstructive sleep apnea (OSA) is an independent risk factor for cardiovascular (CV) morbidity and mortality, but the benefit of continuous positive airway pressure (CPAP) therapy is uncertain. However, most randomized-controlled trials have focused on the role of CPAP in secondary prevention although there is growing evidence of a potential benefit on early CV disease. Weight loss in combination with CPAP may be superior but is difficult to achieve and maintain with conventional measures alone. The aim of this study was to gain insights into the effect of CPAP on early atherosclerotic processes and to compare it to a glucagon-like-peptide (GLP)-1-mediated weight loss regimen in OSA.

**Methods:** We performed a randomized proof-of-concept study (clinicaltrials.gov: NCT04186494) comparing CPAP, a liraglutide-based weight loss regimen (Lir) alone or both in combination for 24 weeks in 30 non-diabetic patients with moderate to severe OSA (50±7 years, 80% males, apnea-hypopnea index [AHI] 50±19/hr, body mass index [BMI] 35.0 ±3 kg/m^2^). Baseline characteristics were similar between groups. Beside extensive evaluation for CV risk factors and endothelial function at baseline and end of study, subjects underwent 18F-fluorodeoxyglucose (FDG)-PET-CT for measurement of aortic wall inflammation (target-to-background ratio [TBR]) and coronary CT angiography (CCTA) for semi- automated coronary plaque analysis.

**Results:** CPAP alone and combination resulted in greater reduction in AHI than Lir alone at 24 weeks (mean difference -45/hr and -43/hr, respectively, vs -12/hr, p<0.05). Both Lir and combination led to significant weight loss of 6±3% and 4±4%, respectively. Despite CPAP resulting in small weight gain, only the CPAP alone group demonstrated a significant decrease in vascular inflammation (aortic wall TBR from 2.03±0.34 to 1.84±0.43, p 0.010) associated with improvement in endothelial function and decrease in C-reactive protein.

Low-attenuation coronary artery plaque volume as marker of unstable plaque also decreased with CPAP (from 571±490 to 334±185mm^3^) and with combination therapy (from 401±145 to 278±126mm^3^) but not with Lir.

**Conclusion:** These data suggest that CPAP therapy, but not GLP-1 mediated weight loss, improves vascular inflammation and reduces low-attenuation coronary artery plaque volume in OSA patients. These novel findings support the benefit of CPAP therapy in modifying early CV disease.

**Clinical Perspective:** *What is new?:* This is the first study comparing standard CPAP therapy to a GLP-1 mediated weight loss regimen in obstructive sleep apnea (OSA). The study utilized 18F-FDG PET CT and artificial intelligence-enabled coronary CT quantification of coronary artery plaque subtypes to determine treatment effects on early atherosclerotic disease processes.

*What are the clinical implications?:* CPAP in contrast to GLP-1-mediated weight loss may improve early atherosclerotic and potentially modifiable disease processes in obstructive sleep apnea (OSA). These data support the benefit of CPAP in the primary prevention of cardiovascular diseases in OSA.

## Introduction

Obstructive sleep apnea (OSA) is a highly prevalent condition conveying a significant public health burden^1,2^. Corroborated by numerous studies it has been identified as a significant and potentially modifiable risk factor for the development and progression of various cardiovascular (CV) diseases, such as coronary artery disease (CAD), hypertension, heart failure and stroke leading to substantial morbidity and mortality^2,3^.

The benefit of continuous positive airway pressure (CPAP) therapy, the treatment of choice for the majority of patients, on CV disease processes, remains uncertain and although impacted by important methodological limitations, secondary prevention trials have failed to demonstrate a significant modifying effect on CV outcomes^4-6^. However, various randomized controlled trials (RCT) and meta-analyses have supported a benefit of CPAP on CV risk factors such as blood pressure^7,8^, endothelial dysfunction^9^, arterial stiffness^10^, and circulating inflammatory markers^11,12^ and thus, CPAP may be particularly beneficial in the early stages of CV disease processes. There is a lack of studies focusing on early atherosclerotic processes in OSA and in the absence of definite primary prevention RCT’s, which are an unrealistic goal in the field, such studies may assist in the guidance of personalized treatments.

Advances in CV imaging techniques may help to fill this gap. The use of positron emission tomography-computed tomographic (PET-CT) imaging with 18F-fluoro2-deoxy-D-glucose (18F-FDG) to measure vascular inflammation is an emerging method of predicting CV disease. In the largest cohort study to date comprising 932 cancer patients without a history of CV disease, aortic wall uptake of 18F-FDG was the strongest predictor of a subsequent vascular event^13^.

Coronary CT angiography (CCTA) is widely used to evaluate CAD and CV risk, traditionally focusing on calcified plaque (CP) and coronary artery calcium scores^14^. However, recent advances in artificial intelligence analysis methods have allowed for semi-automated quantification of non-calcified (NCP) and low-density (LD NCP) plaques, which have been shown to be stronger predictors of future cardiovascular events than calcified plaques^15,16^.

We hypothesized that CPAP improves early CV disease processes. To explore this hypothesis and to inform the design of future large RCT’s, we performed this proof-of- concept prospective RCT comparing CPAP versus a glucagon-like peptide (GLP)-1 (Liraglutide) -based weight loss regimen versus both in combination. This comparison was chosen based on the work of Chirinos et al, who found that a combination of weight loss and CPAP therapy may be superior to either treatment alone across a range of outcomes^17^. We integrated the primary outcome determining early CV disease with various risk markers in relation to inflammation, endothelial function, and blood pressure.

## Methods

### Study Protocol

This was a proof-of-concept, randomized, parallel, 24-week trial comparing the effects of CPAP, a liraglutide-based weight loss regimen (Lir) or both in combination in adults with newly diagnosed moderate to severe OSA (apnea/hypopnea index [AHI]>15/hr). Eligible participants were aged 18-60 years, had a body-mass index (BMI) between 30–40 kg/m^2^, and no history of diabetes, heart failure or unstable cardiovascular disease defined as lack of cardiovascular event within the preceding 3 months. Detailed inclusion and exclusion criteria are provided in Supplementary Table S1 and the trial design with outcome measures is outlined in Supplementary Figure S1. Randomization was conducted using block design. The study was approved by the Ethics Committee of St. Vincent’s University Hospital and registered with clinical trials.gov (NCT04186494). Participants gave written informed consent.

### Interventions

In the CPAP and combined-intervention groups, participants were initiated on CPAP therapy according to the standard protocol in our center. Briefly, patients undergo an education and mask fitting session with a clinical sleep nurse specialist before setup on auto- PAP at home (AirSense 10, ResMed, UK). Adherence to CPAP therapy is monitored remotely and subjects are followed up by the treating clinician and specialist nurse after 4-6 weeks. Pressure settings are adjusted according to the discretion of the treating clinician.

Participants in the Lir and combination groups received education about administration of subcutaneous injections and specific guidance about Liraglutide. In addition, counselling on diet and exercise was provided at 4 and 12 weeks. Advice was based around accepted Irish public health guidelines following the food pyramid recommendations^18^ on portion sizes and food groups and current recommended weekly exercise amounts of 150 minutes of moderate intensity exercise per week^19^. Subjects commenced Liraglutide (Novo Nordisk, Bagsværd, Denmark) 0.6mg once daily at week 1, increasing by 0.6mg weekly until the maximum dose of 3.0mg was achieved, which was then continued for the 24-week intervention. Progress, adverse effects, and safety were monitored routinely at 4 and 12 weeks or in between if required, and serum amylase levels were checked monthly.

### Study Assessments and End Points

All assessments were performed at baseline and at 24 weeks (Supplementary Figure S1). Fasting blood samples for measurement of metabolic, biochemical, and inflammatory parameters were obtained in the morning after overnight polysomnography (PSG). 24-hour ambulatory blood pressure measurement (ABPM) was performed using WatchBP03 devices (Microlife Corp., Taipei, Taiwan). Measurements were taken every 20 minutes during the day and every 30 minutes at night-time. Non-dipping blood pressure status was defined as <10% decline in systolic blood pressure from day to night.

### Polysomnography (PSG)

Attended, in-laboratory PSG was performed using standard techniques (SOMNOmedics, Randersacker, Germany) and analyzed according to the American Academy of Sleep Medicine (AASM) 2.6 scoring criteria^20^. An apnea was defined as a 90% drop in airflow from baseline for >10 seconds. A hypopnea was defined as a drop in airflow of >30% from baseline for >10 seconds, accompanied by an oxygen desaturation of 3% or an arousal. Repeat PSG at 24 weeks was performed on CPAP for those randomized to this treatment.

### Endothelial Function

Evaluation of peripheral endothelial function via the EndoPAT 2000 device (Itamar Medical, Israel) is a validated, FDA-approved method^21^. Endothelial function was expressed as reactive hyperemia index (RHI) calculated as the ratio of the post-occlusion to pre- occlusion peripheral arterial tone (PAT) amplitude of the tested arm, divided by the post- occlusion to pre-occlusion ratio obtained in the control arm^22^. Measurements were performed according to the manufacturer’s instructions with a cut-off value of <1.69 defining endothelial dysfunction.

### 18F-FDG PET-CT

All 18F-FDG PET-CT examinations were performed on a Siemens Biograph mCT PET/CT system (Siemens Healthineers, Erlangen, Germany) in line with prior recommendations^23^ (see Supplementary Methods). PET image analysis to measure aortic wall vascular inflammation was performed in consensus by 2 radiologists blinded to all clinical information, using dedicated PET-CT image viewing software (SyngoVia, Siemens Healthineers, Erlangen, Germany) according to a previously validated methodology^24,25^.

Briefly, sequential circular regions of interest were placed across the aortic wall from the root to the abdominal bifurcation on axial images, with total aorta SUV_max_ calculated as the mean of these individual measurements. A target-to-background ratio (TBR) was calculated by dividing total aorta SUV_max_ by mediastinal blood pool activity (average of three measurements of blood pool SUV_max_ made at the SVC on sequential slices).

### Coronary Computed Tomography Angiography (CCTA) Protocol and Image Analysis

CCTA images were acquired on a 64-slice single-source CT scanner (Gemstone 750 HD, GE, Milwaukee, USA) according to standard CCTA protocols (see Supplementary Methods). Coronary artery calcium (CAC) score was assessed using the Agatston score^26^.

Coronary arteries were visually assessed on a 15-segment basis by a cardiothoracic radiologist. Quantitative assessment of atherosclerotic plaque subtypes was performed using previously validated semiautomatic software (CVi42, Circle Cardiovascular Imaging Inc, Calgary, Canada)^27^ by one observer blinded to all clinical information (see Supplementary Methods). Plaque volumes were measured for the following plaque subtypes: total plaque, calcified plaque (CP), noncalcified plaque (NCP) and low-attenuation plaque (LD NCP) defined by an attenuation of <30 Hounsfield units^16^. CCTA was performed at baseline in all subjects. Ethical approval for follow-up CCTA was only granted for subjects with visible CV disease at baseline due to concern of excess radiation exposure.

### Statistical Analysis

This was an exploratory proof-of-concept study with the purpose of aiding the planning of future larger RCT’s. Therefore, all statistical analysis presented is for the purpose of exploring signals in treatment differences between groups. Comparisons between groups were performed via one-way ANOVA with post-hoc Bonferroni analysis or Chi-Square test for categorical variables. Within-group changes were evaluated by paired T-tests. Univariate Pearson correlation analysis was performed to assess the correlation between baseline and OSA parameters and CV outcome variables. Statistical analysis was carried out using IBM SPSS Statistics Version 27.

## Results

### Baseline characteristics

450 consecutive patients attending our Sleep Centre were screened of whom 53 met the inclusion and exclusion criteria. 23 did not provide consent. As planned, 30 subjects were enrolled and randomized. Baseline characteristics of the study population are provided in Table 1. As expected for a clinical OSA population, there was a predominance of male, middle-aged participants and most had severe OSA with an AHI of 50±19/hour. The groups were similar with regards to demographic, anthropometric and clinical parameters. There was no significant difference between the groups in 24-hour ABPM, glycosylated hemoglobin (HbA1c) and fasting lipids. 9 patients were on statin treatment: 2 for previously established but stable CV disease, and 7 for dyslipidemia as primary prevention.

**Table 1:** Baseline Characteristics of the study population (n = 30). (CPAP = continuous positive airway pressure, BMI = body mass index; BP = blood pressure, AHI = apnea- hypopnea index, HDL = high-density lipoproteins, LDL = low-density lipoproteins, HbA1c =glycosylated hemoglobin. Data are expressed as mean ± SD or n *(%))*

|  | Total | Liraglutide<br>n=10 | CPAP<br>n=10 | Combination<br>n=10 | p-value |
| --- | --- | --- | --- | --- | --- |
| Female | 6 (20) | 2 (20) | 1 (10) | 3 (30) | 0.535 |
| Age (years) | 50±7 | 50±9 | 51±8 | 50±5 | 0.959 |
| BMI (kg/m<sup>2</sup>) | 35.0±3.1 | 35.0±3.1 | 36.0±3.2 | 34.0±3.3 | 0.527 |
| Epworth Sleepiness<br>Scale | 9±5 | 8±6 | 7±4 | 11±5 | 0.245 |
| AHI | 50±19 | 53±20 | 50±21 | 47±16 | 0.816 |
| Hypertension | 7 (23) | 3 (30) | 3 (30) | 1 (10) | 0.475 |
| Ischemic Heart<br>Disease | 2 (7) | 0 (0) | 0 (0) | 2 (20) | 0.137 |
| Statin Use | 9 (30) | 3 (30) | 2 (20) | 4 (40) | 0.621 |
| Anti-Hypertensive<br>Medication Use | 10 (33) | 3 (30) | 3 (30) | 4 (40) | 0.861 |
| Smoking Status: |  |  |  |  |  |
| • Current<br>smoker | 3 (10) | 0 (0) | 2 (20) | 1 (10) | 0.329 |
| • Ex-smoker | 16 (53) | 4 (40) | 6 (60) | 6 (60) | 0.585 |
| • Never-smoker | 11 (37) | 6 (60) | 2 (20) | 3 (30) | 0.155 |
| Smoking Pack Years | 22±10 | 20±14 | 21±9 | 24±10 | 0.812 |
| 24-hr mean systolic<br>BP (mmHg) | 121±12 | 126±13 | 119±10 | 120±12 | 0.359 |
| 24-hr mean diastolic BP (mmHg) | 74±8 | 76±9 | 72±6 | 75±8 | 0.425 |
| Non-dippers | 20 (67) | 7 (70) | 8 (80) | 5 (50) | 0.350 |
| HbA1c (mmol/mol) | 38.4±3.2 | 38.4±3.9 | 37.8±2.6 | 39.0±3.1 | 0.784 |
| Total Cholesterol (mmol/l) | 4.80±0.99 | 5.35±1.17 | 4.68±0.66 | 4.36±0.90 | 0.071 |
| Triglyceride (mmol/l) | 1.88±0.90 | 2.09±0.76 | 1.84±1.00 | 1.72±0.97 | 0.66 |
| HDL Cholesterol (mmol/l) | 1.09±0.23 | 1.13±0.21 | 1.02±0.28 | 1.13±0.21 | 0.458 |
| LDL Cholesterol (mmol/l) | 2.85±0.82 | 3.28±0.89 | 2.83±0.48 | 2.45±0.86 | 0.07 |
| T.Chol/HDL Ratio | 4.53±1.08 | 4.81±1.00 | 4.84±1.10 | 3.95±0.99 | 0.111 |

### Study Progress

All 30 patients completed the trial and are included in the final analysis (Supplementary Figure S2). Of the 10 subjects commenced on CPAP therapy, one patient crossed over to the Liraglutide arm due to intolerance of CPAP therapy. In each of the Liraglutide and combination groups, one subject crossed over to the CPAP arm due to gastrointestinal side effects with Liraglutide. Thus, the final dataset included n=11 in the CPAP group, n=10 in the Liraglutide group and n=9 in the combination group. All groups remained similar in all baseline characteristics (Supplementary table S2). Apart from those having to stop Liraglutide, adherence to the pharmacological treatment was good as reported by the subjects and by monitoring the contents of the pre-filled pens. Serum amylase was monitored monthly as recommended for patients taking Liraglutide, and there was no significant rise during the trial in any subject. Mean CPAP adherence in the CPAP group was 5.8±1.4 hours per night, while adherence in the combination group was 4.7±1.8 hours per night.

### Anthropometrics, blood pressure, fasting lipids and C-reactive protein (CRP)

As demonstrated in Table 2, both Liraglutide alone and in combination with CPAP led to significant weight loss of 6±3% and 4±4%, respectively, and this was accompanied by an improvement in fasting triglyceride levels. In contrast, subjects on CPAP alone showed an increase in body weight although this did not reach statistical significance (2±5%, p=0.092).

**Table 2:** Changes from baseline to 24 weeks intervention. (CPAP = continuous positive airway pressure, CRP = C-reactive protein, HDL = high-density lipoproteins, LDL = low- density lipoproteins, MAP = mean arterial pressure. Data are expressed as mean ± SD. **Bold** indicates significant within-group changes, *p<0.05 vs CPAP alone)

|  | **Liraglutide** | **CPAP** | **Combination** |
| --- | --- | --- | --- |
| Change in Weight (kg) | **-6.17±3.57\*** | 2.30±4.57 | **-3.67±4.22\*** |
| Change in Waist Circumference (cm) | -4.25±6.11 | 0.75±6.45 | **-6.25±2.75** |
| Change in CRP (mmol/l) | -0.89±2.05 | -1.63±2.50 | -0.02±1.64 |
| Change in Total Cholesterol (mmol/l) | **-0.23±0.45** | -0.29±0.50 | -0.28±0.45 |
| Change in Triglycerides (mmol/l) | **-0.49±0.49** | -0.11±0.86 | -0.09±0.81 |
| Change in HDL Cholesterol (mmol/l) | 0.08±0.11 | 0.00±0.14 | 0.08±0.14 |
| Change in LDL Cholesterol (mmol/l) | -0.09±0.41 | -0.25±0.70 | -0.15±0.46 |
| Change in Total Chol/HDL Ratio (mmol/l) | **-0.43±0.34** | -0.25±0.70 | **-0.48±0.49** |
| Change in Systolic Blood Pressure (mmHg) | **-5.51±6.32** | 0.88±5.91 | -2.33±11.66 |
| Change in Diastolic BP (mmHg) | -2.10±3.81 | -0.50±4.75 | -1.89±8.33 |
| Change in MAP (mmHg) | -3.2±4.3 | -0.04±4.4 | -2.01±4.3 |
| Change in Systolic Dip (%) | 0±7 | -4±7 | -7.00±12 |
| Change in Diastolic Nocturnal Dip (%) | -2±9 | **-11±10** | **-15±11** |

No change in 24-hour ABPM was seen in either the CPAP or the combination group, but systolic blood pressure improved in the Liraglutide group (122±8 vs 116±8 mmHg, p = 0.031). At baseline, there were 8 non-dippers in the Liraglutide group, 7 in the CPAP group and 5 in the combination group. There was no significant change in non-dipping status in any treatment group at 24 weeks (Table 2).

CRP levels improved the most in the CPAP group but there were no statistical differences between groups (Table 2).

### OSA parameters

Polysomnographic respiratory variables at baseline and at follow-up are presented in Table 3. As expected, CPAP alone or in combination resulted in effective treatment of OSA. Liraglutide-mediated weight loss was also accompanied by significant improvement in AHI, but the effect was significantly inferior to CPAP, and the AHI remained high at 42±16/hr.

**Table 3:** Baseline and end-of-study obstructive sleep apnea parameters. (CPAP = continuous positive airway pressure, AHI = apnea/hypopnea index, ODI = oxygen desaturation index, TST90 = oxygen saturation time spent below 90% of total sleep time, SpO_2_ = oxygen saturation; Bold indicates p <0.05 for within group changes, *p<0.05 vs Liraglutide)

|  |  | **Liraglutide** | **CPAP** | **Combination** |
| --- | --- | --- | --- | --- |
| AHI | Pre | 54±21 | 48±20 | 48±17 |
|  | Post | 42±16 | **3±3\*** | **5±5\*** |
| ODI | Pre | 45±22 | 40±17 | 43±17 |
|  | Post | 32±19 | **6±11\*** | **5±5\*** |
| TST90 | Pre | 14±12 | 17±17 | 11±11 |
|  | Post | **7±11** | **0±1** | **1±2** |
| Minimum SpO<sub>2</sub> | Pre | 77±10 | 77±6 | 78±7 |
|  | Post | 81±8 | **87±4** | **88±4** |
| Baseline SpO<sub>2</sub> | Pre | 94±1 | 93±2 | 93±1 |
|  | Post | 94±1 | **94±2\*** | 94±1 |

### Endothelial Function

A RHI via the EndoPAT device of <1.69, indicative of endothelial dysfunction, was present in 2 patients in the Lir group, in 7 patients in the CPAP group and in 2 patients in the combination group at baseline (p = 0.036). There was a negative correlation between RHI and overnight baseline oxygen saturation (SpO_2_) (r = -0.527, p = 0.003) and night-time systolic blood pressure (r= -0.360, p = 0.05), but there was no other correlation with OSA variables. Following the treatment protocol, there was a significant improvement in RHI in the CPAP group only (p = 0.048).

### Changes in vascular inflammation evaluated by 18F-FDG PET-CT

Vascular inflammation of the aortic wall measured via PET-CT correlated with the degree of baseline endothelial dysfunction as well as with baseline CRP (Supplementary Table S3). In addition, there was a significant relationship between vascular inflammation and both the magnitude of systolic blood pressure nocturnal dip and nocturnal systolic blood pressure. Furthermore, vascular inflammation correlated significantly with the severity of nocturnal hypoxia as measured via basal oxygen saturation (SpO_2_) (r = -0.499, p = 0.005). There was also a trend towards significant correlation between vascular inflammation and baseline AHI (r=0.342, p=0.064) (Supplementary Table S3).

To elucidate the potential confounding effects of statin treatment on vascular inflammation, we repeated the correlation analysis excluding subjects on statin treatment. Correlations between baseline vascular inflammation and AHI and markers related to the severity of intermittent hypoxia became stronger (Supplementary Table S3) and remained significant after controlling for BMI (AHI: r = 0.482, p = 0.031, oxygen desaturation index [ODI]: r = 0.497 p = 0.026, time spent with SpO_2_<90% of total sleep time [TST90]: r = 0.471, p = 0.036).

Investigating the change in vascular inflammation over 24 weeks, there was a significant reduction in the CPAP group (change in aortic wall TBR from 2.03±0.34 to 1.84±0.43, p 0.010; representative example shown in Figure 1), which was significant when compared to the Liraglutide group (p = 0.028) with a similar trend towards a reduction in inflammation in the combination group. Liraglutide-mediated weight loss alone, however, did not result in a significant change (Figure 2A). The change in vascular inflammation showed a significant negative correlation with change in basal SpO_2_ (r=-0.412, p=0.026) and a close to significant correlation with change in AHI (r=0.337, p=0.074). Again, when patients on statins were excluded, the changes in vascular inflammation in the CPAP and combination groups were enhanced (Figure 2B).

**Figure 1:**
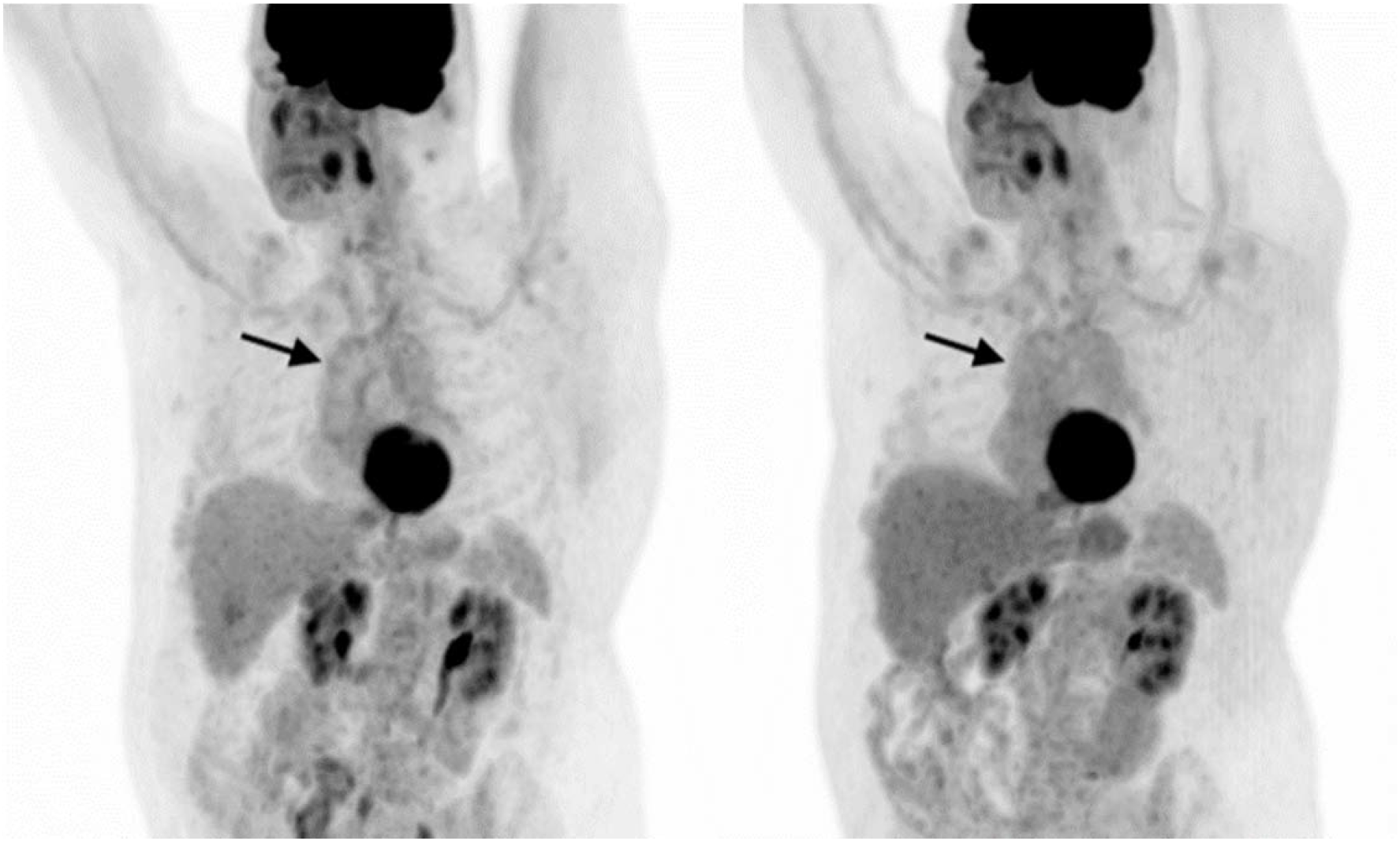
Representative coronal oblique FDG PET maximum intensity projection (MIP) images with mild, diffuse aortic wall uptake in the ascending thoracic aorta (arrows) on the baseline PET (A), which qualitatively resolved on the end of study PET with CPAP alone (B). Aortic wall target-to-background ratio (TBR) was 1.74 on the baseline PET and 1.13 on the end of study scan.

**Figure 2:**
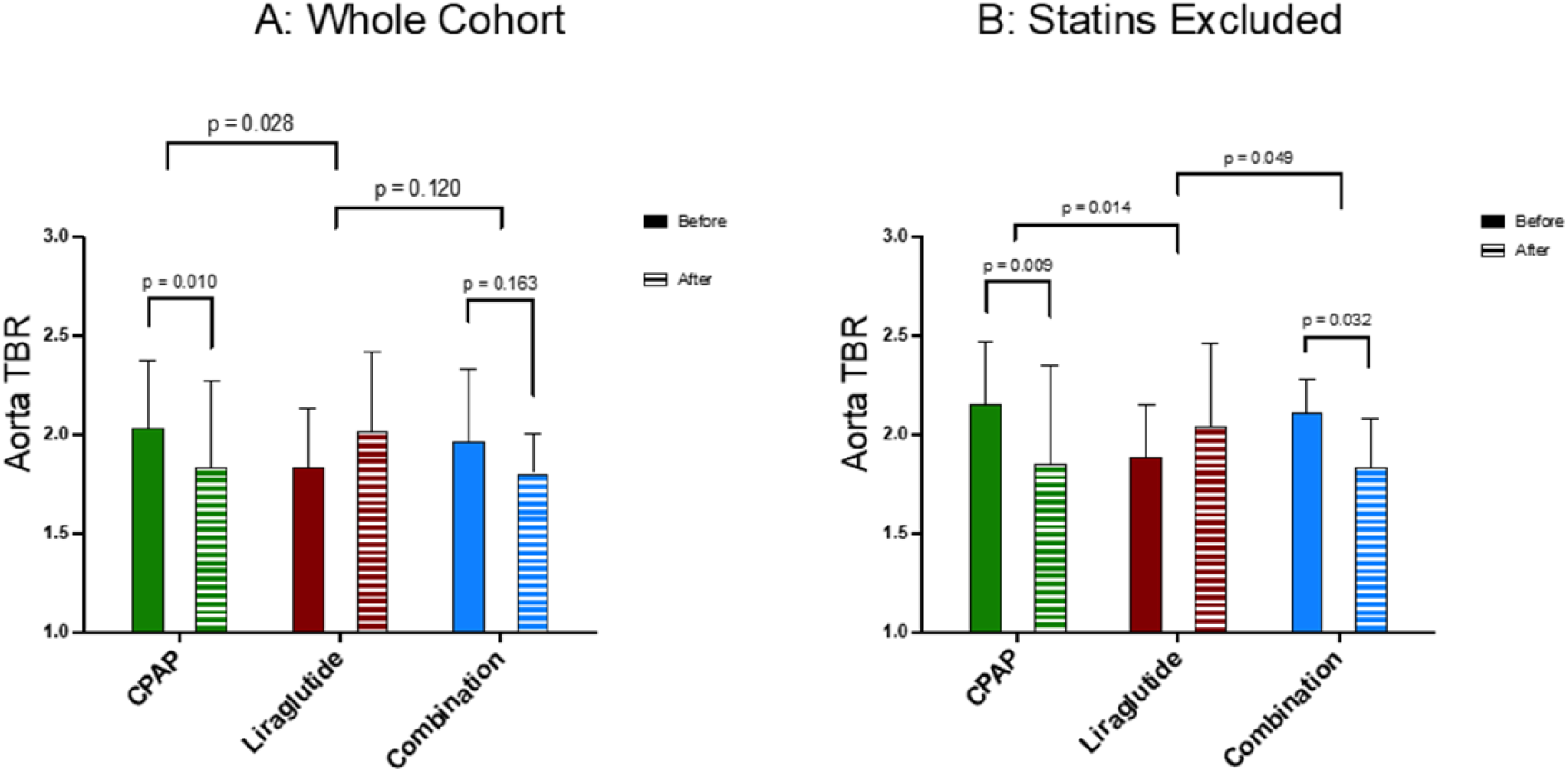
Change in vascular inflammation as measured by aortic wall target-to-background ratio (TBR) in the three treatment groups in A) the whole cohort (n = 30) and B) the cohort with those using statin medication excluded (n = 21). Data represent mean±SD.

### Changes in coronary artery plaque volumes

Visual analysis of baseline CCTA revealed CAD in 17/30 (47%) subjects, of whom 2 had been previously diagnosed. The CAC score in those patients was 65+/-194 with no difference between treatment groups at baseline. Of the 17 subjects with visually identifiable CAD, 11 had single-vessel disease (5 Lir, 3 CPAP, 3 combination), 4 had two-vessel disease (1 Lir, 1 CPAP, 2 combination, and 2 had three-vessel disease (1 CPAP, 1 combination). Quantitative assessment of calcified plaque (CP) volume correlated closely with the visually obtained CAC score (r = 0.767, p <0.001), but there was no relationship between non-calcified (NCP) or low-attenuation (LD NCP) volume with this measure.

At baseline, all plaque subtype volumes were similar between groups. There was a trend towards significant correlation between LD NCP volume and baseline fasting triglyceride levels (r= 0.366, p = 0.051), and this relationship remained similar following adjustment for BMI (r= 0.365, p = 0.056). There were no significant correlations between plaque measures and OSA parameters in the whole cohort, but TST90 correlated with CP volume in the group who did not take statin medication (r = 0.607, p 0.005).

The 17 subjects with visible coronary artery plaque on the baseline scan underwent repeat CCTA after 24 weeks, and those were evenly distributed between groups (6 Lir, 6 CPAP, 5 combination). Although limited given the small group, there was a reduction in LD NCP volume with CPAP (619±532 vs 364±190 mm^3^) and with combination therapy (401±145 vs 278±126 mm^3^), and there was no change with Liraglutide alone (545±335 vs 503±229 mm^3^) (Figure 3). Changes in total and NCP volume followed similar patterns but there were no relevant changes in CP burden with any treatment. Representative images from a study participant of the CPAP alone arm are presented in Figure 4.

**Figure 3:**
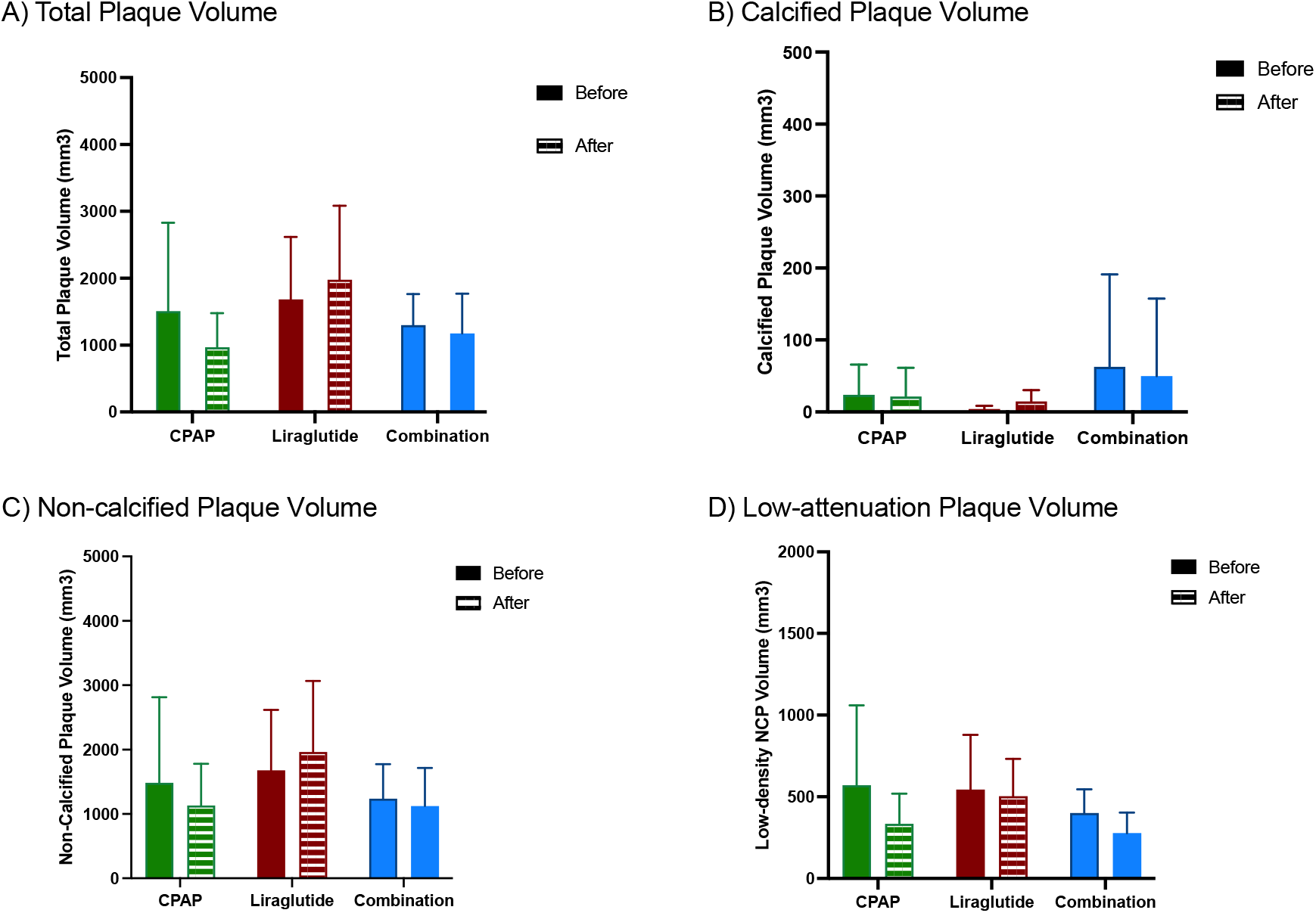
Change in CT coronary artery plaque volumes from baseline to 24-week of intervention. A) total plaque, B) calcified plaque (CP), C) non-calcified plaque (NCP) and D) low-attenuation plaque (LD NCP)

**Figure 4:**
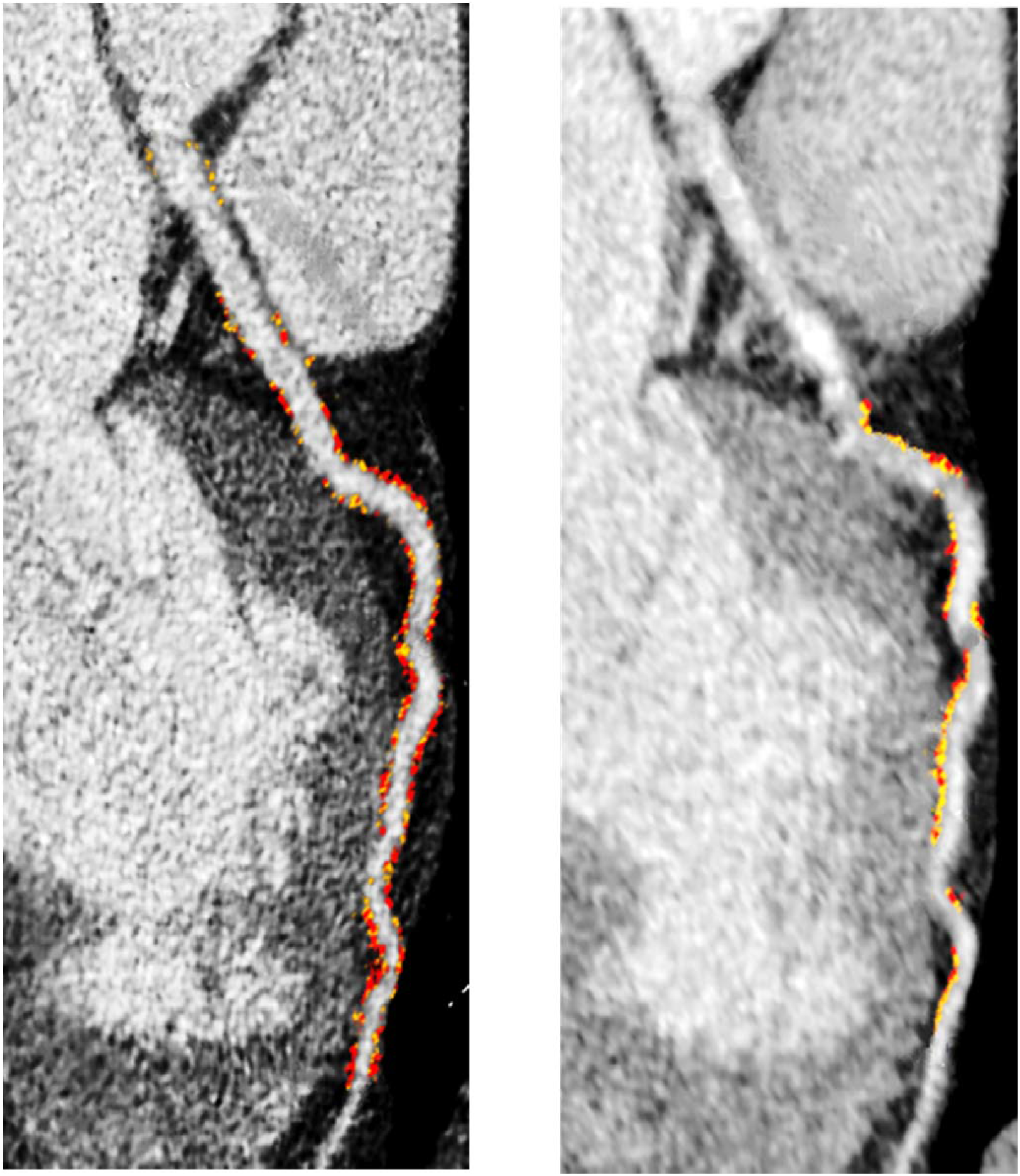
Representative quantitative coronary plaque analysis in a subject assigned to CPAP therapy alone. Images of the left anterior descending coronary artery showed a mixture of non-calcified (yellow color overlay) and low density (red color overlay) plaque subtypes throughout the vessel at baseline imaging (A) (total volume 625mm^3^) and a reduction in both subtypes of 236mm^3^ at 24-week repeat coronary computed tomographic angiography (CCTA) (B).

Evaluating the total group of subjects undergoing repeat CCTA, there was a significant univariate correlation between the reduction in NCP volume and the reduction in TST90 (r = 0.505, p = 0.038), and a trend towards significant correlation with the reduction in AHI and ODI (r = 0.457, p = 0.065 and r = 0.463, p = 0.061, respectively) (Figure 5).

**Figure 5:**
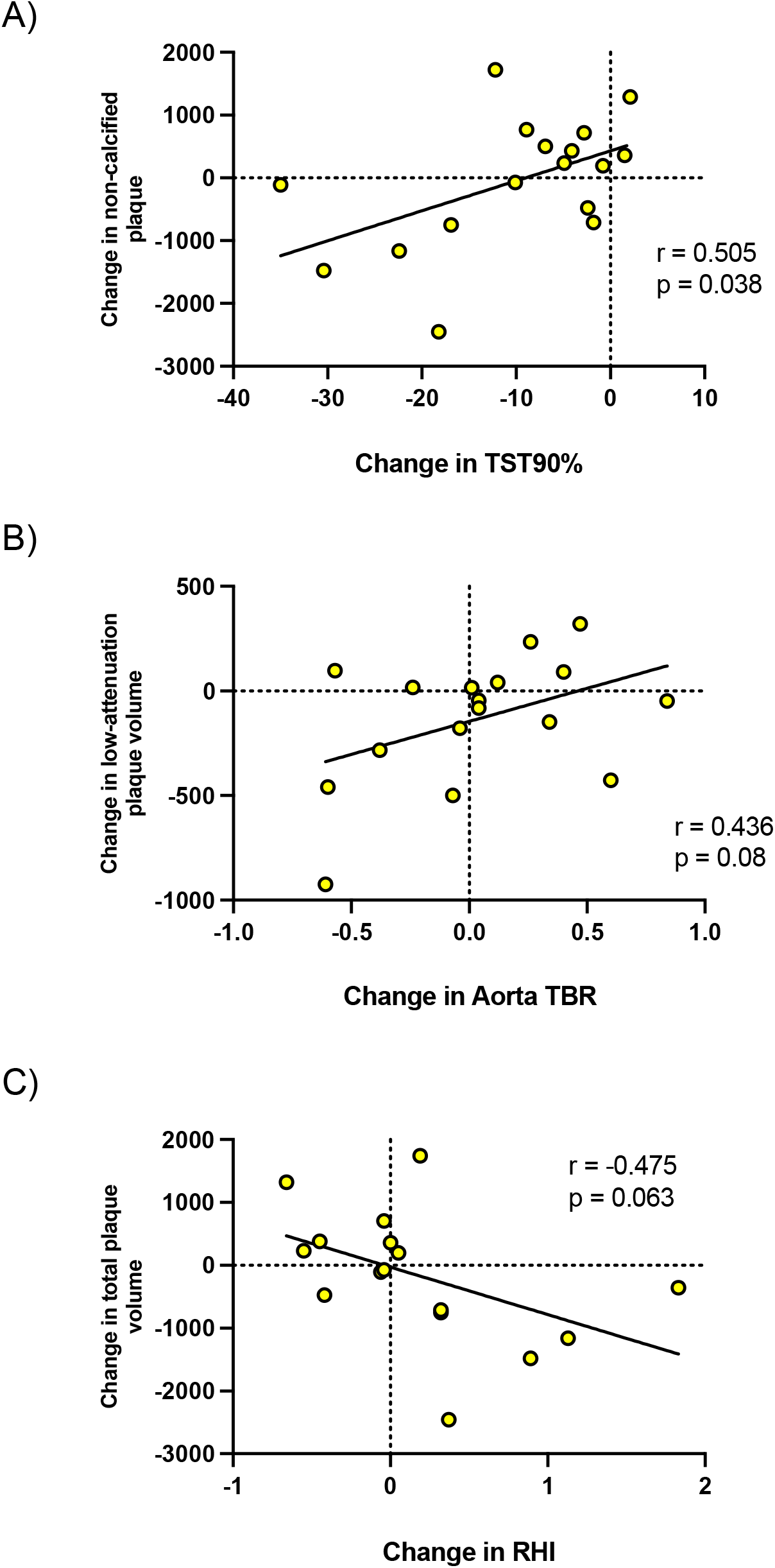
Correlations between change in CCTA plaque measurements and A) change in TST90, B) change in vascular inflammation as measured by aortic wall TBR, C) change in endothelial function as measured by RHI

Furthermore, there was a trend towards significant correlations between the reduction in vascular inflammation and reduction in both NCP and LD NCP volume (r = 0.447, p = 0.072 and r = 0.436, p = 0.080, respectively). Finally, improvement in endothelial function as measured by the RHI showed a close to statistically significant correlation with reduction in total plaque volume, but not with either NCP or LD NCP volume (r = -0.475, p = 0.063).

## Discussion

In this report we demonstrate that CPAP therapy but not a GLP-1-mediated weight loss regimen improves early and potentially modifiable atherosclerotic processes in OSA. As this was a proof-of-concept study with only a small number of participants the statistical differences in improvement in vascular inflammation between the treatment arms are particularly striking and support the benefit of CPAP in the primary prevention of CV disease processes in OSA. The role of CPAP therapy in the treatment of CV diseases and particularly in the prevention of CV events in subjects with OSA has been subject of major controversy. Three randomized controlled trials failed to demonstrate attenuation in the incidence of CV events in cohorts with established CV diseases^4-6^. However, exclusion of patients with severe daytime sleepiness and low adherence to CPAP therapy were substantial limitations and in contrast, analyses from large real-world studies revealed a significant association of CPAP use with lower all-cause mortality^28-30^. Furthermore, several studies and meta-analyses have demonstrated a clinically meaningful improvement with CPAP of various cardiovascular risk factors including blood pressure or endothelial function^8,9^. Importantly, the presence of OSA appears to be particularly detrimental in subjects without prior CV disease^31^. This ties in with an extensive line of evidence showing that intermittent hypoxia, the hallmark feature of obstructive sleep apnea and key pathophysiological factor in CV disease processes in OSA, is a potent triggering factor of inflammatory processes which are also characteristic of early stages of atherosclerosis^32,33^. We and others have previously shown that OSA subjects without CV disease exhibit a low-grade systemic pro-inflammatory milieu which is attenuated by effective CPAP therapy^11,34^. The CPAP effect on aortic inflammation as evaluated by FDG PET-CT, on LD NCP coronary artery plaque burden and on endothelial function found in our present report support these findings and highlight the potential opportunity for early intervention to mitigate CV risk in OSA. Notably, compliance with CPAP was excellent in our cohort and adequate adherence commonly referred to as usage of 4 or more hours per night has emerged as being critical to achieve CV benefit^35^. Our small numbers do not allow further insight into the dose-effect relationship between CPAP and CV processes, but this topic will remain a research priority.

Although adherence and tolerability with Liraglutide were also satisfactory and significant weight loss was achieved accompanied by improvement in triglyceride levels and blood pressure, this did not translate into improvement of early atherosclerotic disease in our study. The general health benefits of weight loss are unquestionable and have also been demonstrated in OSA populations^36-38^. However, most studies have focused on improvement in OSA parameters or metabolic outcomes. In a landmark study, Chirinos and co-workers reported improvement in various parameters with weight loss including insulin sensitivity and blood pressure in obese subjects with obesity and moderate to severe OSA in contrast to CPAP therapy. Of note, there appeared to be an incremental benefit with both treatments combined^17^. However, that study employed intensive weight loss coaching and dietary restriction that are not feasible in real world populations. GLP-1 in combination with lifestyle change is superior in achieving weight loss, compared to lifestyle change alone, and is now widely used as an obesity treatment^39^. The SCALE sleep apnea trial evaluated this pharmacological intervention versus placebo over 32 weeks in moderate to severe OSA subjects who were unwilling or unable to use CPAP and reported a small but statistically significant improvement in AHI, systolic blood pressure and HbA1c^40^. Our study is the first to compare the impact of GLP-1 induced weight loss with CPAP therapy (alone or in combination) and also the first to address the potential impact on early CV disease processes. Weight loss achieved in the Liraglutide group was comparable to that seen in the SCALE sleep apnea trial but was less in the combination group. This was likely due to difficulties for some subjects in managing both treatments concomitantly, also reflected in the lower CPAP compliance in comparison to those on CPAP alone. We used a very pragmatic, easily applicable approach for standard clinical populations. We did not provide specific dietary or exercise advice to patients in the CPAP group reflecting our standard practice. However, as a consequence we cannot rule out a potential added benefit on CV disease with combined CPAP and Liraglutide. Furthermore, continuous weight loss beyond our 24-week timepoint might potentially relate into more noticeable improvements but this remains hypothetical at this stage. Nonetheless, our data support a direct contribution of OSA to the process of atherosclerosis and the significant correlations of oximetry parameters with our CV outcomes reinforces the pivotal role of intermittent hypoxia as triggering factor. The relationship between OSA and CV disease was particularly strong in subjects not on statin therapy. Statin medication is the cornerstone therapy to attenuate CV disease progression and their effects are not solely mediated by their lipid-lowering properties but also through pleiotropic effects reducing vascular inflammation and promoting atherosclerotic plaque stability^41,42^. Statin use among OSA patients has been consistently reported to be low ^43,44^ and studies assessing the benefits of statins in OSA populations have been conflicting. In a French multicenter double- blind RCT endothelial function remained unaffected although statins improved blood pressure and lipid profile^45^. In contrast, a recent report demonstrated reduction of complement-mediated pro-inflammatory effects with restoration of endothelial protection^46^. Our study was not designed to assess this aspect but supports the need for further research into the role of statins in OSA-mediated CV disease either in combination with CPAP or as primary preventive strategy alone.

A particular strength of our study is the evaluation of early and potentially reversible atherosclerotic disease using state-of-the art imaging techniques. This is the first time that 18F-FDG PET-CT has been used in an OSA population. Vascular inflammation on 18F-FDG PET-CT has been shown to predict the formation of future atherosclerotic plaques and moreover, to correlate with subsequent vascular events^13,47^. Importantly, the reduction in aortic inflammation by CPAP therapy in our study is at least of the magnitude seen with statin therapy^48^ and thus, is likely of clinical relevance. In contrast to PET-CT assessment of atherosclerosis, CCTA is well established in routine clinical practice and increased calcified coronary artery plaque burden on CT calcium scoring is predicative of future coronary events^49,50^. We reported previously an increased plaque burden in otherwise healthy OSA subjects versus matched controls^51^ and data from the Multi-Ethnic Study of Atherosclerosis (MESA) cohort demonstrated an independent association between OSA severity and progression of calcified coronary artery plaques over an 8-year period^52^. However, calcified plaques represent a more stable form of atherosclerosis less likely to rupture and cause myocardial infarction. Histologically, a lipid-rich necrotic core is an important characteristic associated with plaque rupture^53^ and those can be accurately identified on CCTA as low-attenuation noncalcified plaque and accurately quantified through semi-automatic analysis. Accordingly, low-attenuation plaque has been demonstrated to be a better predictor of myocardial infarction than other traditionally used markers including coronary artery stenosis^54^. In a large cross-sectional study of 692 subjects, OSA severity correlated strongly with CP and NCP burden and subjects with moderate to severe OSA were more likely to contain a LD NCP component than controls^55^. Unfortunately, we were not able to repeat CCTA’s in all our subjects following their interventions due to the ethical restrictions regarding radiation exposure but nonetheless with our small numbers, there was a noticeable trend in reduction with CPAP in LD NCP volume only, supporting the potential importance of effective OSA treatment on the early pro-inflammatory phase of atherosclerosis.

These data have been derived from a proof-of concept study and thus, have inherent limitations including low numbers, an open-label design and a focus on a selected, well-characterized population. In addition, in line with other RCT’s in the field, we excluded subjects with severe daytime sleepiness who may benefit most from CPAP.

Despite these limitations, our data provide sufficient evidence to mandate the design and delivery of larger-scale studies to define the role of CPAP in attenuating CV risk as primary prevention in subjects with OSA. There is also a clear need to integrate novel radiological surrogate markers of cardiovascular disease with clinical endpoints.

## Data Availability

The data that support the findings of this study are not openly available due to reasons of sensitivity and are available from the corresponding author upon reasonable request.

## Non-standard Abbreviations and Acronyms

OSA: obstructive sleep apnea
CV: cardiovascular
CAD: coronary artery disease
CPAP: continuous positive airway pressure
RCT: randomized controlled trial
PET-CT: positron emission tomography-computed tomography
FDG: fluoro2-deoxy-D-glucose
CCTA: coronary computed tomographic angiography
CP: calcified plaque
NCP: non-calcified plaque
LD: low density
GLP: glucagon like peptide
AHI: apnea/hypopnea index
BMI: body mass index
ABPM: ambulatory blood pressure measurement
PSG: polysomnpgraphy
AASM: American Academy of Sleep Medicine
RHI: reactive hyperemia index
PAT: peripheral arterial tone
TBR: target-to-background ratio
CAC: coronary artery calcium
HbA1c: glycosylated hemoglobin
SpO_2_: oxygen saturation
ODI: oxygen desaturation index
TST90: time spent with SpO_2_<90% of total sleep time

## Acknowledgement

We are grateful to all the patients and their families for participating in this study and to all staff members of the Sleep Laboratory at St. Vincent’s University Hospital for their help and support.

## Supplementary Material

Supplemental Methods

Tables S1-S3

Figure S1-S2

